# Population age and household structures shape transmission dynamics of emerging infectious diseases: a longitudinal microsimulation approach

**DOI:** 10.1101/2023.06.05.23290874

**Authors:** Signe Møgelmose, Laurens Vijnck, Frank Neven, Karel Neels, Philippe Beutels, Niel Hens

## Abstract

Host population demographics and patterns of host-to-host interactions are important drivers of heterogeneity in infectious disease transmission. To improve our understanding of how population structures and changes therein influence disease transmission dynamics at the individual and population level, we model a dynamic age-and household-structured population using longitudinal microdata drawn from Belgian census and population registers. At different points in time, we simulate the spread of a close-contact infectious disease and vary the age profiles of infectiousness and susceptibility to reflect specific infections (e.g. influenza and SARS-CoV-2) using a two-level mixing model, which distinguishes between exposure to infection in the household and exposure in the community. We find a strong relationship between age and household structures, which, in combination with social mixing patterns and epidemiological parameters, shape the spread of an emerging infection. Disease transmission in the adult population in particular is explained by differential household compositions and not just household size. Moreover, we highlight how demographic processes alter population structures in an ageing population and how these in turn affect disease transmission dynamics across population groups.

## 1. Background

Host population demographics and patterns of host-to-host interactions are important drivers of heterogeneity in infectious disease transmission. To describe the dynamics of infections transmitted via close contact interactions, particular attention has been given to social mixing patterns, which can be captured by demographic structures. The frequency and pattern of social mixing with relevance for the spread of close-contact infectious diseases are highly dependent on age. Children, teenagers and young adults have more contacts and are disproportionately more likely to mix with people of their own age than with adults older than 25 years. Adults also display age-assortative mixing, but their average contact frequency is lower and their contacts are less concentrated in their own age group than those of youngsters [1,2]. Consequently, age patterns are seen in susceptibility and exposure to many pathogens [3]. Additionally, changes in the immune system throughout the life course can add to these age-specific differences. Children tend to be more susceptible to infections given that their immune system is still maturing, while in older adults, the ageing of the immune system (immunosenescence) may increase their susceptibility to infection and to more severe disease upon infection [4]. Likewise, the infectiousness of infected individuals may also vary by age [5,6].

Population structures beyond age further add to the heterogeneity in social mixing patterns and in disease transmission dynamics. Due to the high frequency, long duration and intimacy of within-household contacts, household transmission constitutes a substantial risk factor in infectious disease dynamics [7,8]. Moreover, households often contain people from different generations (e.g. parents and children) belonging to different subpopulations outside the household, which, for example, can facilitate the spread of an infection from schools to workplaces [9]. Consequently, age-and household-structured models of infectious disease transmission with social mixing have proven highly valuable for modelling the transmission of close-contact infectious diseases [e.g. 10–15].

Still, it remains challenging to model an age-and household-structured population, and in particular the changes therein, in a well-founded and feasible manner. Detailed household data is usually unavailable, which often makes it necessary to re-create households by relying on probability matching and/or to make simplifying assumptions, like limiting to specific household sizes or types (e.g. nuclear families). Less common household constellations such as long-term care facilities (LTCFs) or multi-generational households are often disregarded, although they may be important for disease transmission [16]. Moreover, only few studies incorporate evolving age and household structures in the host population or consider multiple populations with different compositions [e.g. 11,17–20]. While demographic change can be reasonable to disregard when the period under consideration is short, it may be necessary to allow for changing population structures when investigating disease transmission dynamics and control strategies in different populations or over a longer time frame (i.e. years or decades depending on the population, infection and research question), where demographic changes become more apparent. Moreover, a thorough understanding of the relationship between disease transmission dynamics and host population structures, as well as the demographic processes underlying these structures, may be important for assessing how future demographic changes potentially could affect transmission dynamics.

Demographic change has in many countries led to an increasing median age of the population (population ageing), which has become a global phenomenon [21]. Many of the most developed countries have been ageing for decades as a result of declining fertility rates and increasing life expectancy, and the ageing of the large generations born in the mid-twentieth century is currently causing a temporary acceleration of population ageing [22].

We investigate how emerging infections are spreading in a relatively old (i.e. high median age), and still ageing, host population. We use longitudinal microdata drawn from Belgian census and population registers, including individual-level information on age, sex, household membership and kinship links. The data feeds into a demographic microsimulation, which includes dynamic demographic processes for fertility, mortality, migration and household transitions, making it possible to model the Belgian population over time with evolving age and household structures.

We subsequently combine the demographic microsimulation with a two-level mixing model, which distinguishes between exposure to infection in the household and exposure in the community at large. We base contact networks within households on empirical data, rather than making the common assumption of random mixing. We simulate the spread of an emerging close-contact infectious disease in 2020, 2030, 2040 and 2050, which allows sufficient time for noticeable changes in age and household structures to emerge. Furthermore, we vary the age profiles of infectiousness and susceptibility to reflect specific infections, including influenza and SARS-CoV-2.

We aim to explore how the relationship between age and household structures affects disease transmission dynamics of an epidemic at the individual and the population level. Moreover, we show how demographic processes alter the population structures in an ageing population and investigate how this affects the transmission dynamics across population groups.

The paper is organised as follows: In Section 2, we give specific details on the demographic microsimulation, including the demographic data and processes considered. Similarly, we describe the disease transmission model with two levels of mixing along with the model parameters. Section 3 presents the population structures and changes therein and documents the disease incidence by age and household composition. Furthermore, the impact of epidemiological heterogeneities within the population is visualized in a scenario analysis. Finally, in Section 4, we discuss the results as well as the strengths and limitations of the study.

## 2. Methods

### 2.1 Demographic microsimulation

We developed a demographic microsimulation to simulate the Belgian population from 2011 to 2051. The initial population in the microsimulation is based on the Belgian census from January 1^st^ 2011, from which we drew a sample of about 10% of the total population. For each individual, we have information on their date of birth, sex, coded ID of parents, birth trajectory (parity and date of most recent birth if applicable), household ID and household position (e.g. in union, child, single parent). Thus, individuals can be linked to each other through household membership and kinship.

In each time step (i.e. day), individuals can enter and leave the population as a result of births, deaths and migration. Moreover, the household position of an individual may change and transitions between households or the creation of a new household can take place. Finally, ageing takes place at the end of the time step and the population is updated accordingly. The probability of a demographic event taking place varies by individual characteristics, including age, sex and household position, and changes over time except for the household transition rates.

We assume that mortality, fertility and migration levels in the microsimulation resemble the observed and projected rates by the Belgian Statistical Office (StatBel) and the Belgian Federal Planning Bureau (FPB). This implies below-replacement level fertility (a total fertility rate below 2.1 [23]) and continuous improvements in longevity, especially for males [24,25]. Consequently, the population will continue ageing, with implications for the household structures.

The demographic data, model and source code are described in detail in the supplementary material.

### 2.2 Disease transmission model

Disease outbreaks take place in the simulated population every ten years since demographic change is a slow process and substantial effects of population ageing will only emerge after several years. In 2020, 2030, 2040 and 2050, ten randomly chosen individuals become infected, in an otherwise fully susceptible population, on January 1^st^ of each respective year. We consider an infectious disease transmitted via close contact, which can be represented by a Susceptible-Infectious-Recovered (SIR) model and consider several scenarios for age-specific susceptibility and infectiousness. The probability of becoming infected, and thus moving from the susceptible to infectious state, is calculated using a two-level mixing model, where an individual can acquire infection as a result of interactions with an infected household member (local contact) or an infected individual in the general population (global contact) [26]. For each parameter setting in the two-level mixing model, we run 30 simulations, but in the analysis we disregard those where an outbreak never takes place.

#### 2.2.1 Within-and between-household interactions

We use a contact matrix to estimate social interactions (i.e. a proxy for an at-risk event at which infection can be transmitted) between non-household members in the general population. The contact matrix is based on social contact data collected in a survey in Belgium in 2010-2011 [2] and made available through the SOCRATES data tool [27] (Figure S1). Contacts between household members were excluded, as these are captured by the household level of the model, but contacts taking place in the household with non-household members were included. Additionally, supplementary professional contacts (SPC) were excluded.

To model interactions among household members, we construct a household contact network for each household in the population. Specifically, each household member is represented by a vertex, and a link between two vertices indicates a contact between those two household members. The links are constructed using an exponential random graph model developed by Krivitsky et al. [28], which was fitted to data from two contact surveys conducted in Belgium in 2010-2011 [2,9]. The model accounts, amongst other things, for the type of household and the age-sex composition. In each time step (i.e. day), we apply the fitted model from Krivitsky et al. [28] to the households in the simulated population and simulate who comes into contact with whom within each household. The mean network density (the number of links in a household relative to the number of possible links [29]) by household size and type are shown in Figure S2.

#### 2.2.2 Risk of infection

Each susceptible individual *i* acquires infection at time *t* with probability *p*_i_(*t*):

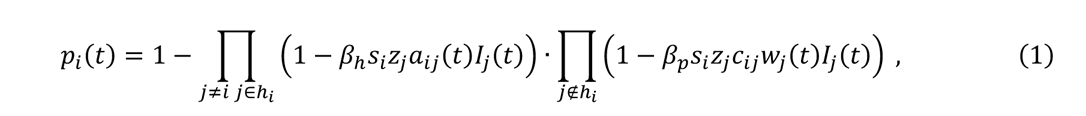

where ℎ_i_ denotes the household of individual *i* and the parameters *²*_h_ and *²*_p_ represent the probabilities of disease transmission given contact between a susceptible and infectious individual within the household and in the general population, respectively. We vary the transmission parameters to reflect different settings (e.g. high vs. low household transmission). The relative susceptibility and infectiousness given the ages of individual *i* and *j* are represented by *S*_i_ and *Z*_j_, respectively, while *I*_j_(t) takes the value one if individual *j* is infected at time *t* and zero otherwise. The contact network in household ℎ_i_ is represented by an adjacency matrix *A* of which the element *a*_ij_(*t*) equals one if household members *i* and *j* come into contact with each other at time *t*, and zero otherwise. A new adjacency matrix is generated in each time step.

The social contact matrix from Figure S1 is represented by *C* and the element *c*_ij_ corresponds to the rate at which *i* and *j* come into contact with each other in the general population given their age. We assume disease transmission in the general population to be frequency-dependent, meaning that the number of effective contacts made by each person remains unchanged as the population grows. Thus, to keep the age-specific contacts constant over time, the changing age composition needs to be taken into account. For that reason, the population size of each age group used to compute the contact rates from Hoang et al. (2021) is divided by the size of the corresponding age group in the simulated population at time *t.* The vector *w* contains the resulting weights, which are updated as the age composition changes. In each time step, the probability of infection (1) is computed for all susceptible individuals in the population and their disease state is updated accordingly.

#### 2.2.3 Infectious period

We assume that the infectious period follows a gamma distribution with a mean of 3.8 days and a standard deviation of 2 days, reflecting the infectious period for influenza [7,30,31]. For each newly infected individual, a value is drawn from the distribution. An infected individual recovers and obtains immunity when the infectious period has passed.

#### 2.2.4 Age-specific susceptibility and infectiousness

We consider different scenarios for age-specific susceptibility and infectiousness to reflect specific infections [4–6,32–34], including influenza and SARS-CoV-2, but also to assess the role different population groups play in the spread of an infection. The age-specific susceptibility and infectiousness scenarios are shown in Figure 1 in relative terms, meaning that a value below one corresponds to reduced susceptibility or infectiousness and a value larger than one implies increased susceptibility or infectiousness for individuals in the given age group. Susceptibility is age-dependent (different from one) in scenario S1-S4, while infectiousness is age-dependent in scenario I1-I4. We compare the different scenarios to a baseline case where infectiousness and susceptibility are equal across all ages.

**Figure 1:**
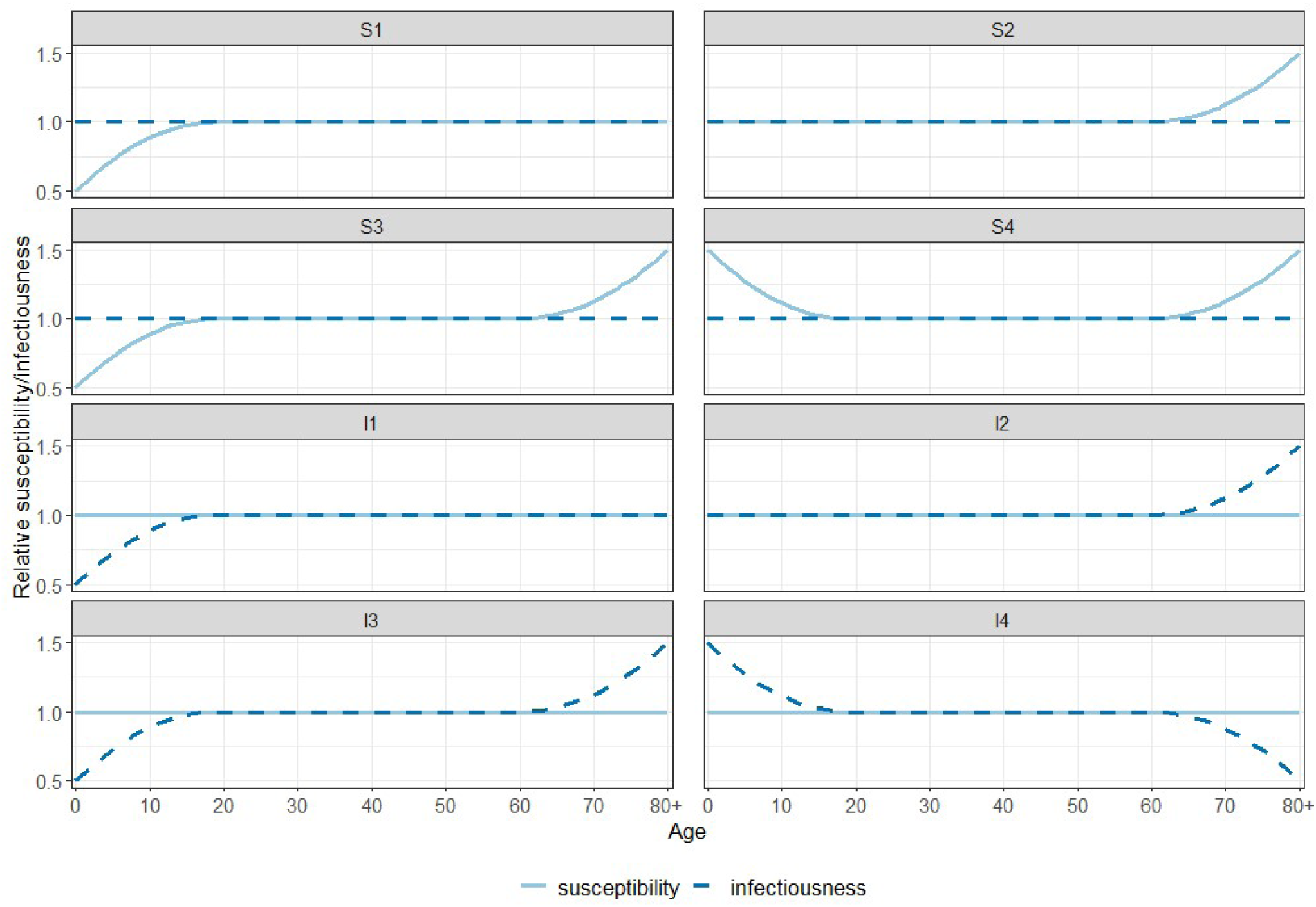
Age-specific susceptibility and infectiousness scenarios.

## 3. Results

### 3.1 Population dynamics

Age and household size are closely connected as seen in Figure 2. Children and adolescents most often live with their parent(s) and siblings, meaning that households of size three and larger are most common at young ages, which implies a similar pattern in the parental age groups (e.g. ages 30-55). The average household size starts to decrease in late adolescence, as children leave the parental household, and increases again from the late twenties with the entry into parenthood. Again, a similar pattern is seen in the parental generation (e.g. age 50+), but with a continuous decrease in mean household size until the mid-seventies, when widowhood and the need for LTCFs become more prevalent. Consequently, single-person households and very large households (i.e. LTCFs) become more frequent in the oldest age groups.

**Figure 2:**
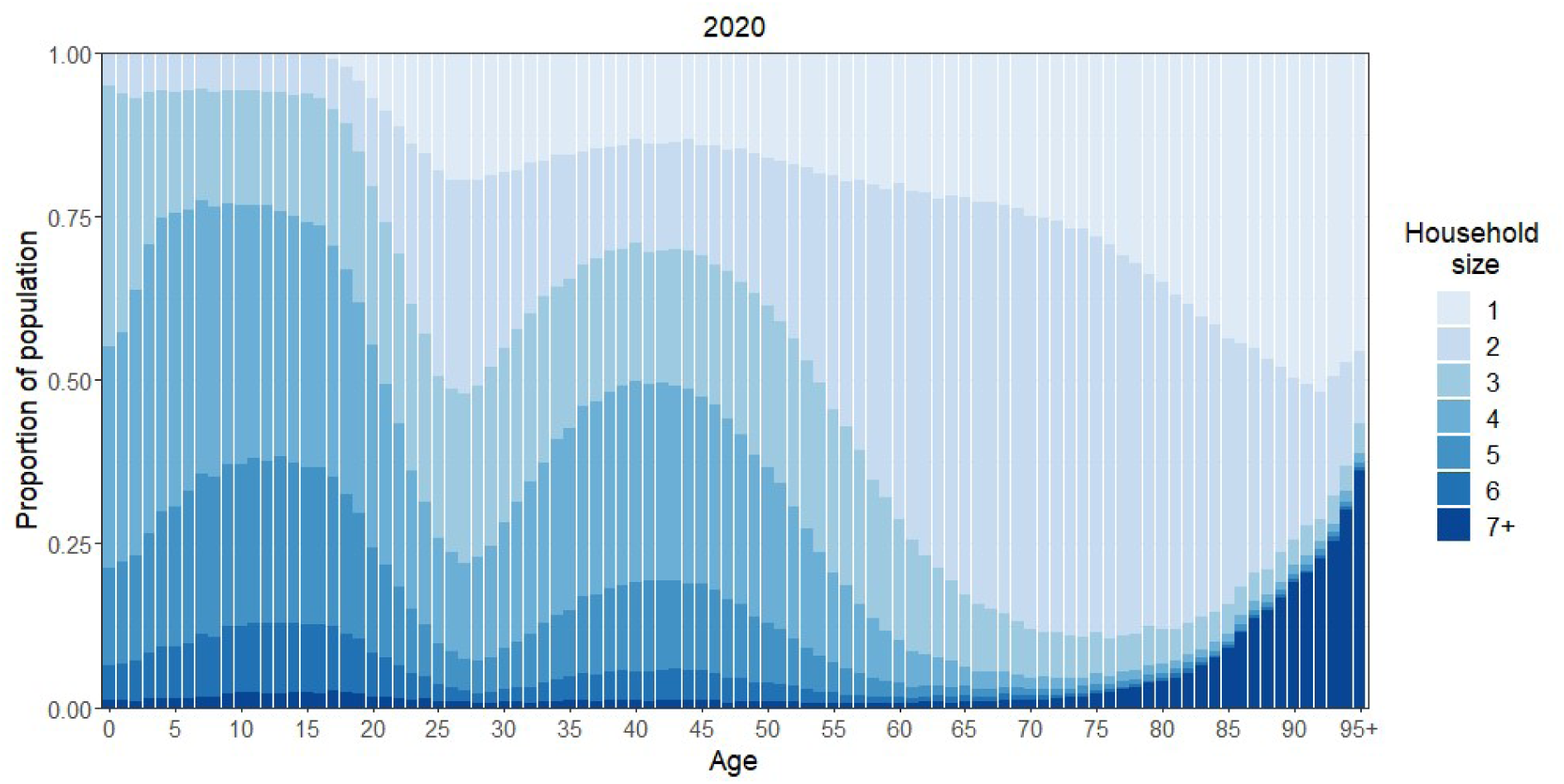
Household size distribution by age group of simulated population in 2020.

The simulated population is ageing between 2020 and 2050, meaning that the elderly population makes up an increasing proportion of the total population as seen in Figure 3 (left panel). This is the result of past long-term trends of declining fertility and increasing longevity, which continue to a certain degree in the simulation period. Moreover, the ageing of the large generations born in the mid-twentieth century causes a temporary acceleration of the population ageing. The changes in the age distribution are also reflected in the household size distribution as seen in Figure 3 (right panel). The proportion of the population living in a single-person household increases as the population ages, since the proportion of people living alone is higher in the older age groups. The proportion living in households of size two is increasing from 2020 to 2040, which is mainly due to the increased survival of elderly males in a union.

**Figure 3:**
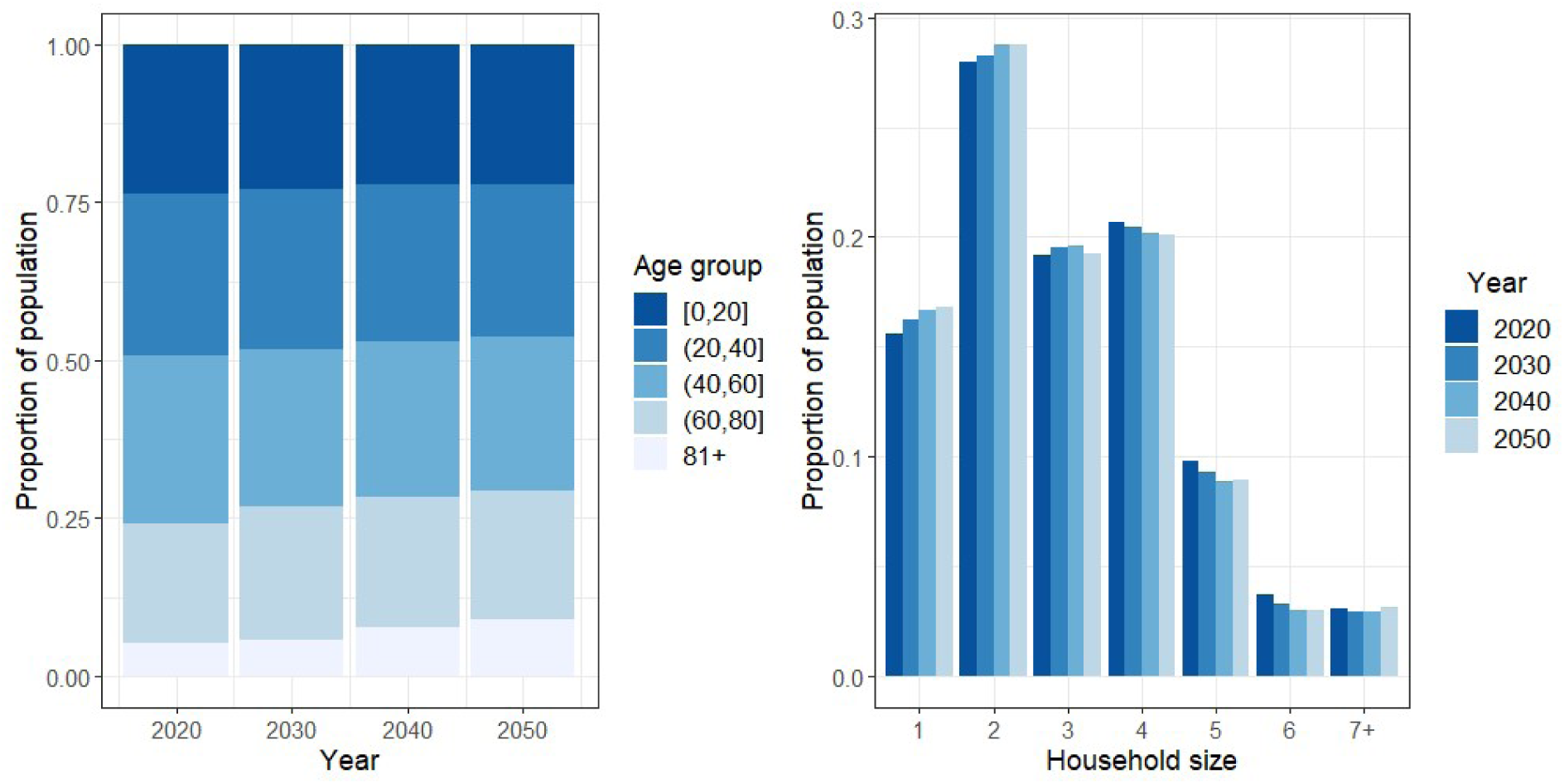
Age distribution (left) and household size distribution (right) of the simulated population.

While an increasing share of the population lives in small households of size one to three, the proportion of larger households of size four to six is decreasing. Households of those sizes are to a large extent occupied by nuclear families in the age range of 0-50 years (see Figure S3-S4), which makes up a decreasing proportion of the simulated population from 2020 to 2050. The proportion of the population living in households of size seven and larger remains quite stable during the simulation period, but it is the result of two opposite trends. The proportion of parents with a large number of children, and therefore with a large household, is decreasing, while people living in LTCFs (i.e. elderly population), make up an increasing proportion of the population.

The age-specific household size distributions slightly change over time (see Figure S3-S4). The average household size for children and their parents (i.e. younger than 50 years) is decreasing. This is a consequence of single parent families becoming more prevalent and a decreasing total fertility rate (the number of live births a woman would have born at the end of the reproductive age span, if she throughout her lifetime were subject to the age-specific fertility rates observed in a given year [23]) prior to 2020, followed by a slow, but not full, recovery (see Figure S5). Meanwhile, the average household size in age group 50-70 is increasing as the likelihood that their household accommodates (adult) children increases, due to past changes in the timing of childbearing. A change in the household size distribution is also seen in the elderly population as a result of improvements in longevity. Consequently, single-person households and collective households (i.e. LTCFs) are increasingly being replaced by two-person households.

### 3.2 Disease transmission dynamics

#### 3.2.1 Attack rate

The proportion of the population that contracts the infection during an outbreak (the attack rate) responds to changes in the transmission parameters (*²*_h_ and *²*_p_ in equation (1)) in a non-linear pattern (see Figure S6). The relative increase in the attack rate diminishes as the transmission probabilities increase, especially when the transmission probability given contact within the household increases. Potential household infections are limited by the size of the household and more than 15% of the simulated population live in single-person households. Thus, as the household transmission probability keeps increasing, everyone with household members will eventually become infected, and only individuals living in single-person households may escape the infection.

This pattern is also seen in the threshold parameter *R*_∗_, which is the product of the expected number of infections within the households (*μ*) and the expected number of secondary cases in the general population (*R*_G_) (see section *Threshold parameter R*_∗_ in supplementary materials). *R*_G_ follows a more or less linear trend while the increase in *μ* becomes smaller as the household transmission probability increases (see Figure S7-S9).

The attack rates in each susceptibility and infectiousness scenario deviate from the baseline scenario since some population groups face an increased or decreased risk of acquiring or transmitting the infection given the specific scenario. However, the differences between the scenarios diminish as the transmission probabilities increase.

#### 3.2.2 Age-and household-specific transmission dynamics: Baseline scenario

Some population groups are more likely to get infected than others, also when discarding age-specific differences in susceptibility and infectiousness (baseline scenario) as seen in Figure 4. The proportion of children and adolescents getting infected is larger than that of any other age group. The adult population also faces relatively large attack rates, which decrease from age 50 onwards. This reflects the age pattern in social contacts outside the household (see Figure S1).

**Figure 4:**
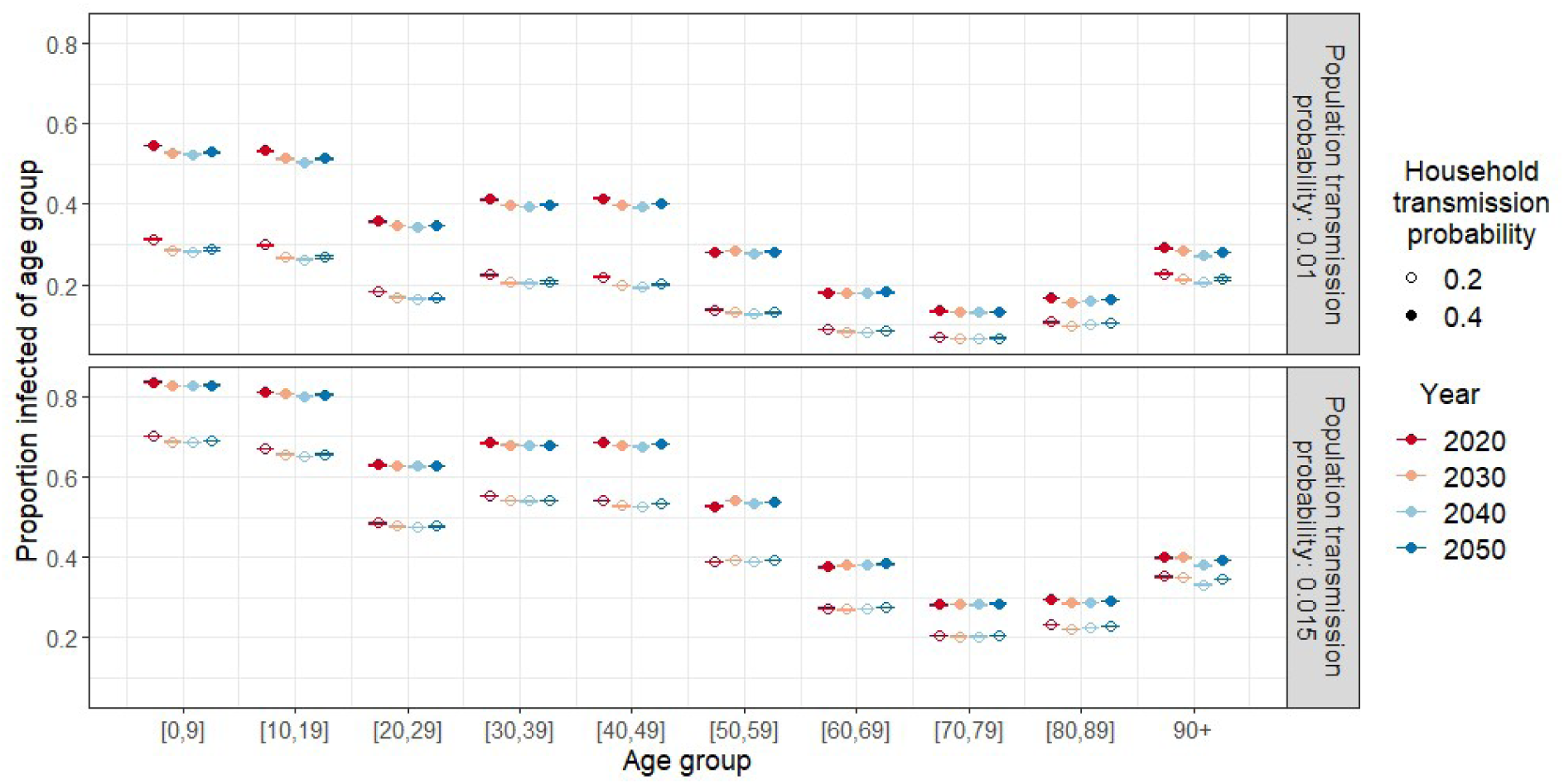
Mean age-specific attack rate with 95% confidence interval for varying transmission parameters (*²*_h_ : filled vs. open circle, *²*_p_: upper vs. lower panel) in the baseline scenario (susceptibility and infectiousness are equal across age).

Nevertheless, social mixing in the general population alone cannot explain the age distribution in the attack rates. The age group 20-29, for example, has more contacts in the general population than the age group 30-49, yet lower attack rates. This is due to the difference in household composition of these age groups. Individuals in their 20s are much more likely to live in households of size one or two, and thereby have a lower number of possible household contacts than people of age 40-49, who often live in larger households as seen in Figure 2.

Moreover, the household members of the two groups tend to differ in case of a larger household. People aged 30-49 living in a household of size three or larger often have young children living with them, while the 20-to 29-year-olds are more likely to live together with their parents or unrelated adults in a house-sharing arrangement (see Figure S4). The mean density of the contact network is higher in the first household constellation than in the latter because of the presence of young children (see Figure S2). Finally, children and teenagers are more likely to bring an infection into the household given their high number of contacts in the general community, including schools, thus putting the parents at an increased risk (assuming that susceptibility and infectiousness are independent of age).

The attack rates in the oldest age groups can also only be explained by considering household structure. The attack rate is higher in age group 90+ than in 80-89, despite both age groups having identical social contact rates outside the household. From the age of 80 onwards, small households of size one and two are increasingly replaced by very large households (see Figure 2), such as LTCFs, which are associated with a high risk of disease transmission, as seen in Figure 5.

**Figure 5:**
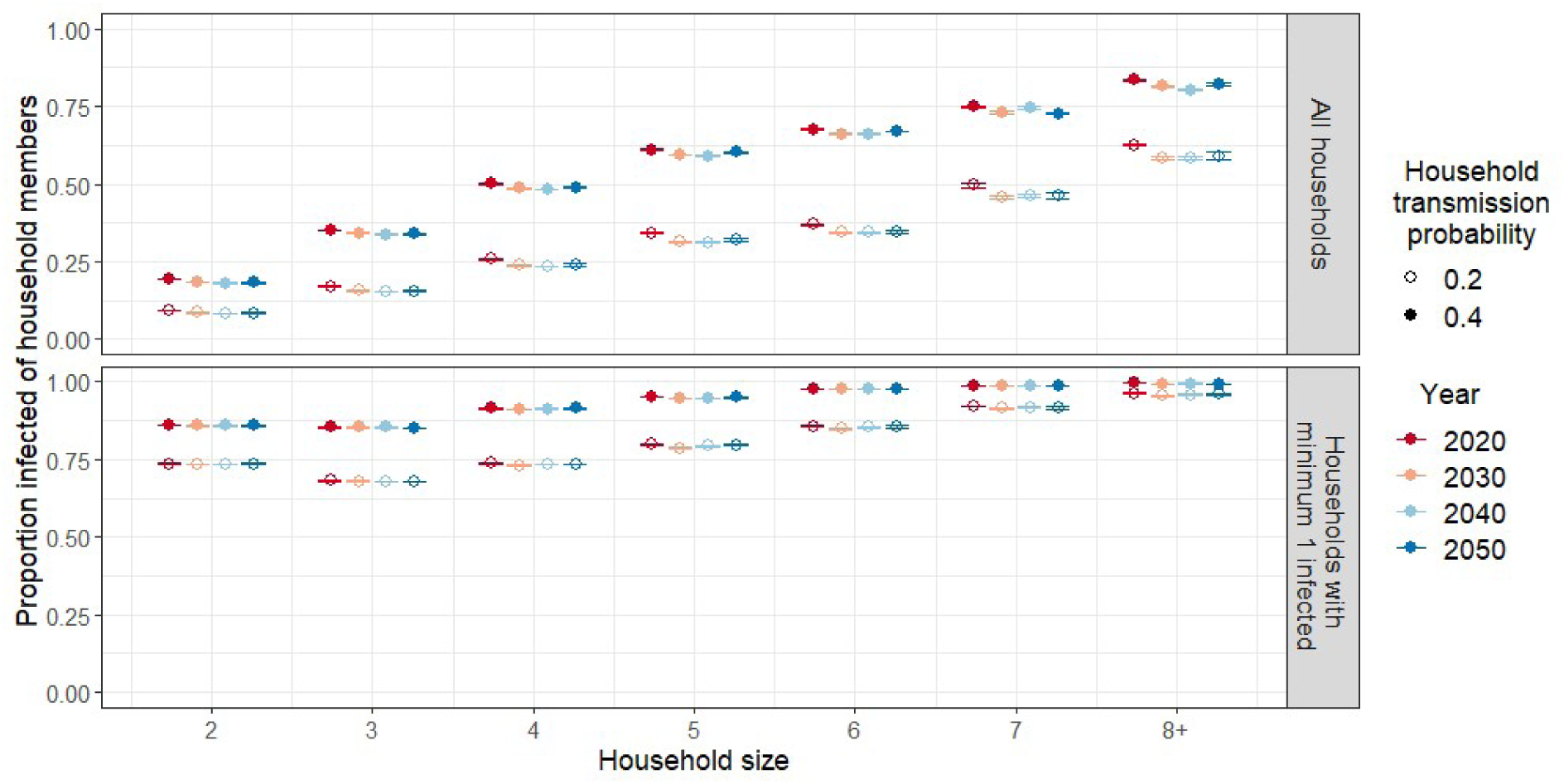
Mean proportion of household members getting infected by household size in baseline scenario with population transmission parameter of 0.01. Upper panel: Estimate based on all households. Lower panel: Estimate based on households with minimum one infected individual.

The household-specific attack rate (i.e. the proportion of household members acquiring infection) increases by household size (Figure 5 upper panel). This result is a combination of how likely an infection is to enter the household and how easily it spreads within that household. The number of individuals that can bring an infection into the household increases with the household size, however, the likelihood of it happening also depends on the social contact patterns of each household member. The infection is more likely to spread to households with at least one child younger than 13 years than to households of the same size without children, because children have a relatively large number of contacts in the community (see Figure S10).

After the infection has entered a household, the further spread is affected by the household size and composition. The within-household transmission is visualised in the lower panel of Figure 5, as the estimates are limited to households with at least one infected individual. In that case, the differences across household sizes are substantially smaller and the mean proportion of household members getting infected even decreases from household size two to three. The decrease, however, is only observed for households without a young child. The presence of a child in the household affects the within-household transmission across all household sizes of less than eight, as young children tend to have more contacts with their household members (i.e. parents, siblings) than teenagers and young adults do with theirs (see Figure S11).

#### 3.2.3 Effect of demographic change on transmission dynamics: Baseline scenario

The proportion of the population acquiring the infection during an outbreak is decreasing from 2020 to 2040 in all scenarios but the trends stabilises or reverses between 2040 and 2050 (see Figure S6). This is the result of changing household structures and population ageing. The elderly population, which over time makes up an increasing proportion of the population, has relatively few contacts on average since the majority lives in small households and has few contacts in the general population. Consequently, the elderly population has a lower risk of acquiring and transmitting an infection than younger age groups.

Additionally, the changing household compositions in the population younger than 50 years of age resulting from low fertility levels and an increasing proportion of single parent families, decreases their risk of infection over time, with implications for the overall incidence. Meanwhile, the proportion infected of age group 50-70 remains more or less stable, despite an increasing proportion living in households larger than size two. Finally, improved longevity implies that the elderly population (especially females) becomes more likely to live with their partner than alone or in LTCFs, which affects the incidence in the oldest age group (90+).

The relationship between risk of infection and household size persists as the population is ageing, but the proportion of infected household members is decreasing over time across all household sizes (Figure 5 upper panel). Meanwhile, the within-household transmission remains stable over time (Figure 5 lower panel).

#### 3.2.4 Effect of age-specific infectiousness and susceptibility

We further investigate the role different age groups play in the spread of an infectious disease by comparing the age-specific attack rates across the susceptibility and infectiousness scenarios using the baseline scenario from Figure 4 as a reference (see Figure 6). This is visualised for varying household transmission probabilities (light vs. dark bar) and population transmission probabilities (upper vs. lower panel).

**Figure 6:**
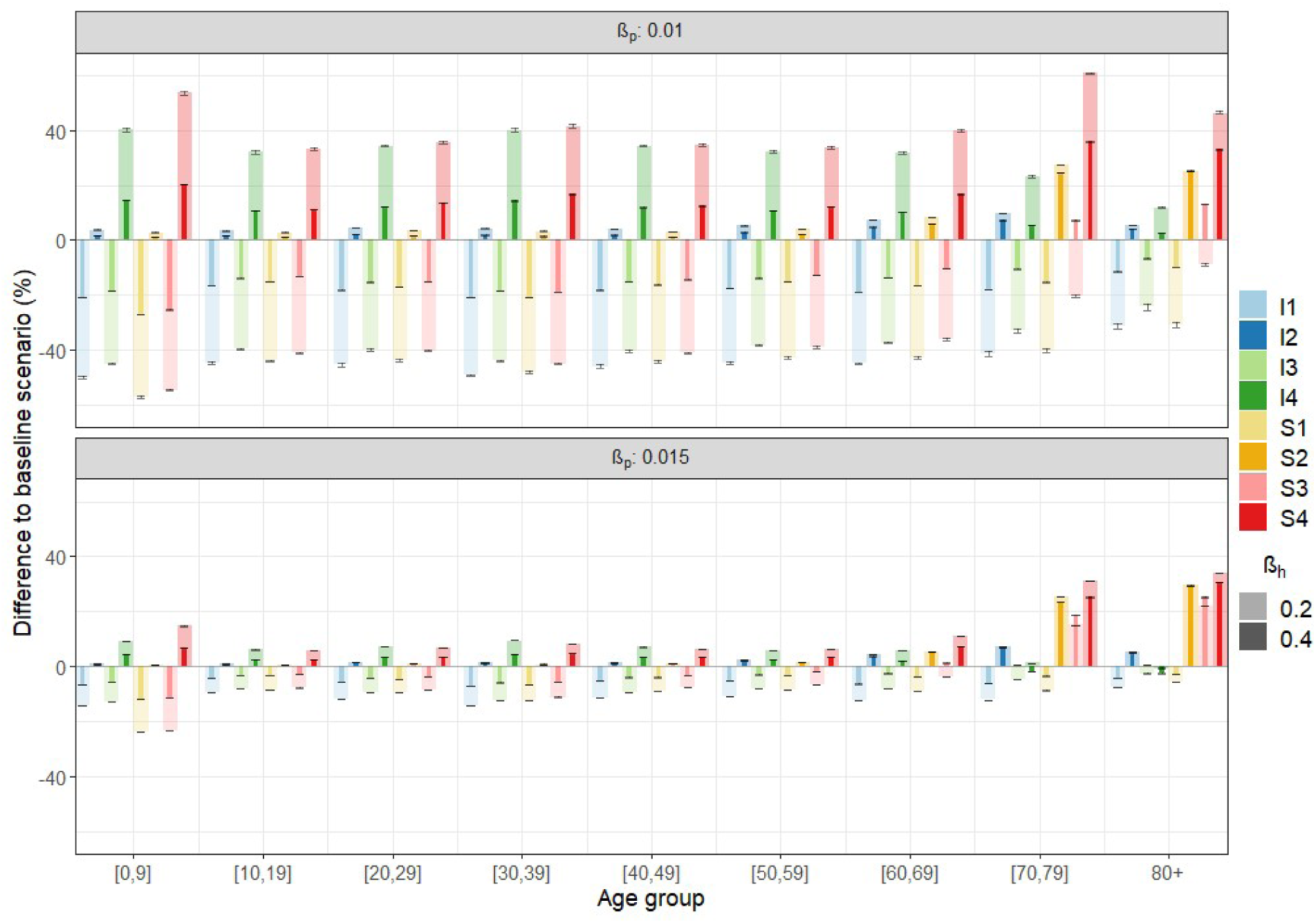
Percentage difference in age-specific attack rate compared to baseline (equal susceptibility and infectiousness across all ages) for varying susceptibility/infectiousness scenarios, population transmission probabilities (upper vs. lower panel) and household transmission probabilities (light vs. dark bars). Simulation year 2020.

Differences from the baseline attack rate are not only seen in the age groups with modified susceptibility or infectiousness, but also in the rest of the population to varying degrees. The susceptibility and infectiousness of children affect all population groups, and the parental generation in particular (e.g. age group 30-39), to a markedly larger degree than changes in the susceptibility and infectiousness of the elderly population. In scenario S1 (I1), children have a relatively low susceptibility (infectiousness) which affects all other age groups substantially, while the relatively high susceptibility (infectiousness) from age 65 onwards in scenario S2 (I2) has a much smaller effect on the incidence in other age groups.

This is also seen by comparing scenarios S1 and S3 (I1 and I3), where the attack rates below the age of 60 do not differ substantially, despite the increased susceptibility (infectiousness) of the elderly population in the latter scenario. Even the elderly population itself is only somewhat affected by changes in their infectiousness. However, it applies to children as well as the elderly, that a change in their infectiousness only has a slightly larger impact on the incidence in the rest of the population compared to the same change in susceptibility.

As the household transmission probability (light vs. dark bar in Figure 6) and population transmission probability (upper vs. lower panel in Figure 6) increase, the differences between the scenarios diminish, except for the age group 70 and older in scenario S3. In scenario S3, children have relatively low susceptibility while the old age groups have high susceptibility. The incidence in the total population remains below the baseline scenario (Figure S6) because the high susceptibility in the elderly cannot compensate for the low susceptibility in children. However, as the transmission probabilities increase, a larger proportion of households with elderly individuals are reached (upper vs. lower panel) and more individuals within the households become infected (light vs. dark bar) due to the high susceptibility. In some cases, the difference to the baseline scenario goes from negative to positive values. This is only observed in the oldest age groups due to the combination of low initial attack rates and high susceptibility.

The relative differences between the different scenarios do not change over time, but in some cases the difference to the baseline scenario changes slightly (Figure S12-S13).

## 4. Discussion

An understanding of demographic structures in the host population and how these influence disease transmission can be important for determining which population subgroups are most at risk and most effective to target in an intervention [35]. Moreover, an understanding of the demographic processes underlying the population structures may be important for assessing how future demographic changes potentially could affect transmission dynamics.

Using longitudinal microdata drawn from Belgian census and population registers, we model a host population with evolving age and household structures using microsimulation and illustrate how population ageing and changing household dynamics may further unfold in the next decades. We combine the demographic microsimulation with a two-level mixing model and illustrate a strong link between age and household structures and the implications thereof for the risk of infection for different population subgroups during an epidemic. Additionally, we quantify the potential impact of changing age and household structures on disease transmission.

The age structures in the social contact patterns are mirrored in the age-specific attack rates as the youngest age groups, with the most community contacts, have the highest risk of infection. The attack rates in the adult population, however, can only be explained by considering the differences in household compositions between young adults, middle-aged adults and older adults, which are related to the timing of demographic events. Young adults in their 20s face a relatively low risk of infection on average, despite a high number of community contacts, because many live in small households (e.g. with a partner or alone). From their late 20s onwards, the proportion of young adults living in households with small children increases along with the attack rate. Thus, the age at entry into parenthood determines when the risk of infection for young adults starts increasing again. The child will eventually have a relatively high number of contacts outside the household (e.g. in day-care, school) as well as frequent contact with the parent(s) within the household, making the risk of infection high compared to other population groups. These relationships change to some degree in the different scenarios for age-specific infectiousness and susceptibility.

The incidence in families with children and/or adolescents decreased during the simulation period as a consequence of changing household compositions. In the decade prior to the first outbreak simulations, 2010-2019, the total fertility rate decreased followed by a slow, but not full, recovery during the remaining simulation period, similarly to observed and projected rates by StatBel and FPB [25]. Changing fertility levels combined with an increase in single parent families affected the household compositions and indirectly disease transmission. This also affects disease transmission in other population groups since families with (school-age) children are important drivers in an epidemic.

As the children grow up and, in most cases, eventually leave the parental household, the number of household contacts of the now middle-aged parental generation is decreasing again, often in combination with decreasing community contacts, leading to lower attack rates. However, we found that the proportion of middle-aged people with (adult) children living in their household is increasing during the simulation. This change is related to the postponement of parenthood since the probability of leaving the parental household is assumed to be constant over time. Parents are increasingly older when the last child leaves the parental household because the average age at childbirth was increasing prior to 2011 when our simulation begins [36]. Several years later, these past fertility trends affect the household composition of the middle-aged population and indirectly their risk of infection. The increasing household size was expected to increase the risk of infection in the middle-aged population, but the effect is more or less counterbalanced by the decrease in the overall incidence induced by the changing household compositions in the younger age groups.

It should be noted that the relative distribution of births by the age of the mother is only slightly changing in the first decades of the simulation and eventually stabilises. However, the average age at childbirth in Belgium has increased since 2011 and this is expected to continue in the future, to some degree [25]. Hence, the average age at childbirth, and indirectly the age at which children leave the parental household, is likely to increase more than in our microsimulation.

The risk of infection in the elderly population was also found to be highly dependent on their household constellation. Community contacts are decreasing with age and a large proportion of old people live alone or only with their partner, which minimises the number of occasions where transmission of a close-contact infection can take place. However, from the age of 80 onwards, the proportion of the population living in LTCFs, which tend to be very large households, increases considerably and so does the risk of infection. Despite a low contact network density in large households, the high number of residents in LTCFs increases the risk of an infection entering the household and it can persist in the household over a long period, making it more difficult to escape transmission. Consequently, the age pattern in the attack rates in the elderly population is shaped by the proportion living in collective households.

During the simulation period, collective households and single-person households in the population older than 80 years of age are increasingly being replaced by two-person households, as a result of improved longevity, particularly of males. We assume that the sex differential in mortality decreases during the simulation period as a result of larger improvements in men’s mortality than in that of women. This implies that an increasing proportion of elderly women are living with a partner instead of alone, which intuitively should increase their risk of infection. However, the probability of moving to LTCFs, which are very large households associated with a high risk of infection, is substantially higher for elderly people living alone than for those living with a partner. Consequently, gradually fewer elderly people move to LTCFs and therefore incur a substantially lower risk of infection.

The future household constellations and mortality of the elderly population are, like all other demographic processes, associated with uncertainty [37]. However, the sex differential in mortality has been decreasing and this is considered likely to continue, to some extent, in the future. The resulting changes in the household structures of the elderly population seen in our microsimulation are in agreement with existing studies of past and future trends in the living arrangements and mortality of older adults [38,39].

In addition to social contact patterns, age and household structures, we also explored how epidemiological heterogeneities within the population may influence the spread of an infection. We incorporated different scenarios for susceptibility to infection when exposed and infectiousness when infected according to age. As anticipated given the social contact patterns and household structures, the susceptibility and infectiousness of children and adolescents were highly influential for the disease transmission in the whole population and in the parental generation in particular. Changes to these epidemiological parameters in the elderly population clearly affected that age group, but exerted much less influence on other age groups.

In some settings, however, the elderly population is affected differently by changes in the transmission parameters than the rest of the population. This was the case in a scenario (S3) with relatively low susceptibility for children and relatively high susceptibility for the elderly, which to some extent can be considered to resemble SARS-CoV-2 infection [33,40–42]. As the probability of transmission given an effective contact increases, the underestimation of the attack rate in the elderly population when omitting age-dependent susceptibility (baseline scenario) increases, while it decreases in the rest of the population, and in all other scenarios.

In a low transmission setting, many old people escape infection due to their limited number of contacts within and outside the household. However, as the transmission probabilities increase, a proportionately larger share of households with elderly people are reached by the infection and more individuals within the households become infected due to the elevated susceptibility. Thus, the impact of epidemiological heterogeneities (e.g. age-specific susceptibility) is not only dependent on the transmission parameters but also on other heterogeneities in the population, including social contact patterns and household structures.

Overall, we find a strong relationship between age and household structures at the individual and population level, which, in combination with social mixing patterns and epidemiological parameters, shape the spread of an emerging infection. Disease transmission in the adult population in particular is explained by differential household compositions. Moreover, we highlight how demographic processes alter population structures with differential impact on the disease transmission dynamics across population groups. Nevertheless, our study faces several limitations with regard to demographic modelling as well as infectious disease modelling.

We recognise that the demographic processes in the microsimulation are simplifications of highly complex processes and that the inherent uncertainty in population projections preferably is described in the form of probability distributions [43,44]. Moreover, the sensitivity in the association between demographic and epidemiological outcomes could have been explored. However, expanding our demographic microsimulation is not considered necessary to fulfill the aim of this paper, which is to document the impact of population structures and the changes therein on the spread of an emerging infection.

In our model of infectious disease transmission, we assume a fully susceptible population, restricting our study to emerging infections. Expanding the study to endemic infectious diseases requires not only information on age-specific patterns of prior immunity but also information on how immunity is distributed in households. Alternatively, a population at an endemic disease equilibrium can be generated, for example by simulating disease transmission over a long period of time before the actual analysis [11,13]. However, this would require a rather complex technique and/or detailed (historical) demographic data. Moreover, if the fertility and mortality schedules remain constant while generating an endemic disease equilibrium, the population eventually acquires the age distribution of the stable population associated with those underlying schedules of vital rates, which may not resemble the population of interest [23].

Another limitation in our model of disease transmission concerns the social contact patterns. If SPCs had been included in the social contact matrix, the incidence in the population of working age would have been slightly higher, however, the relationships found between population groups would remain. Additionally, we assume that the contact patterns in the community and within the households remain constant over time despite the demographic changes. Methods for adjusting social contact matrices to evolving demographic structures have been proposed, but these are not based on empirical evidence for how contact patterns behave over longer time frames as population structures evolve [45,46]. A comparison of two social contact surveys in Belgium five years apart suggests that the contact rates can be assumed stable, but the demographic change in this period is of course limited [2].

## Data Availability

The input data of this study are available from Statics Belgium but restrictions apply to the availability of these data, which were used under license for the current study, and so are not publicly available. A detailed description of the data and source code are available from the GitHub repository: https://github.com/signemoegelmose/demographic_microsimulation_EXTERNAL

## Acknowledgements

The authors are grateful for the contributions made by Andrea Torneri, Pietro Coletti, Pavel N. Krivitsky and Nicholas Geard.

