## Supplementary material for "Population age and household structures shape transmission dynamics of emerging infectious diseases: a longitudinal microsimulation approach"

### Demographic microsimulation

A detailed description of the demographic data and model implementation as well as source code are available from the GitHub repository:

https://github.com/signemoegelmose/demographic_microsimulation_EXTERNAL

### Social contact matrix and household network density

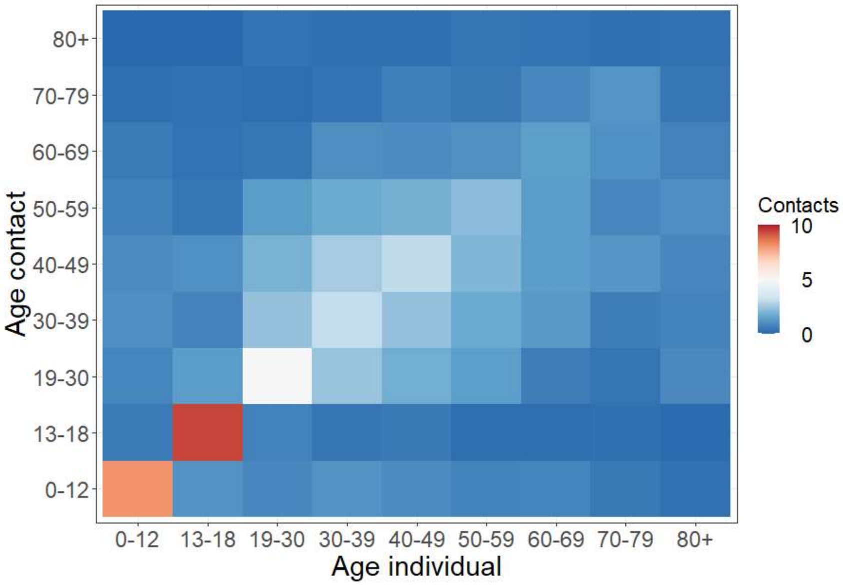

*Figure S1: Age-specific social contacts excluding contacts with household members and excluding supplementary professional contacts (Hoang et al., 2021; Willem et al., 2020).*

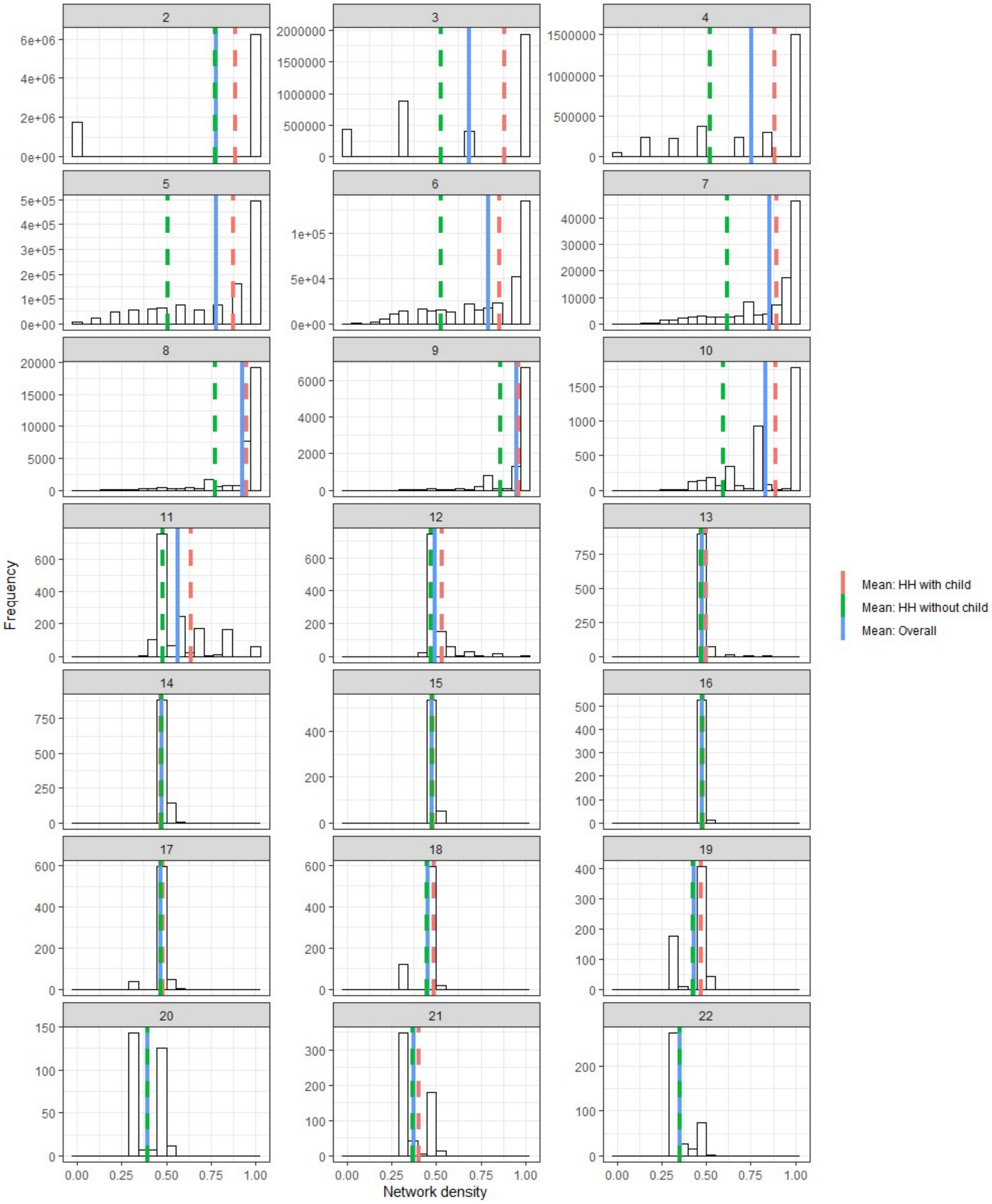

*Figure S2: Histograms of household network densities by household size (HH: household, child: age < 13).*

### Household size and position by age groups

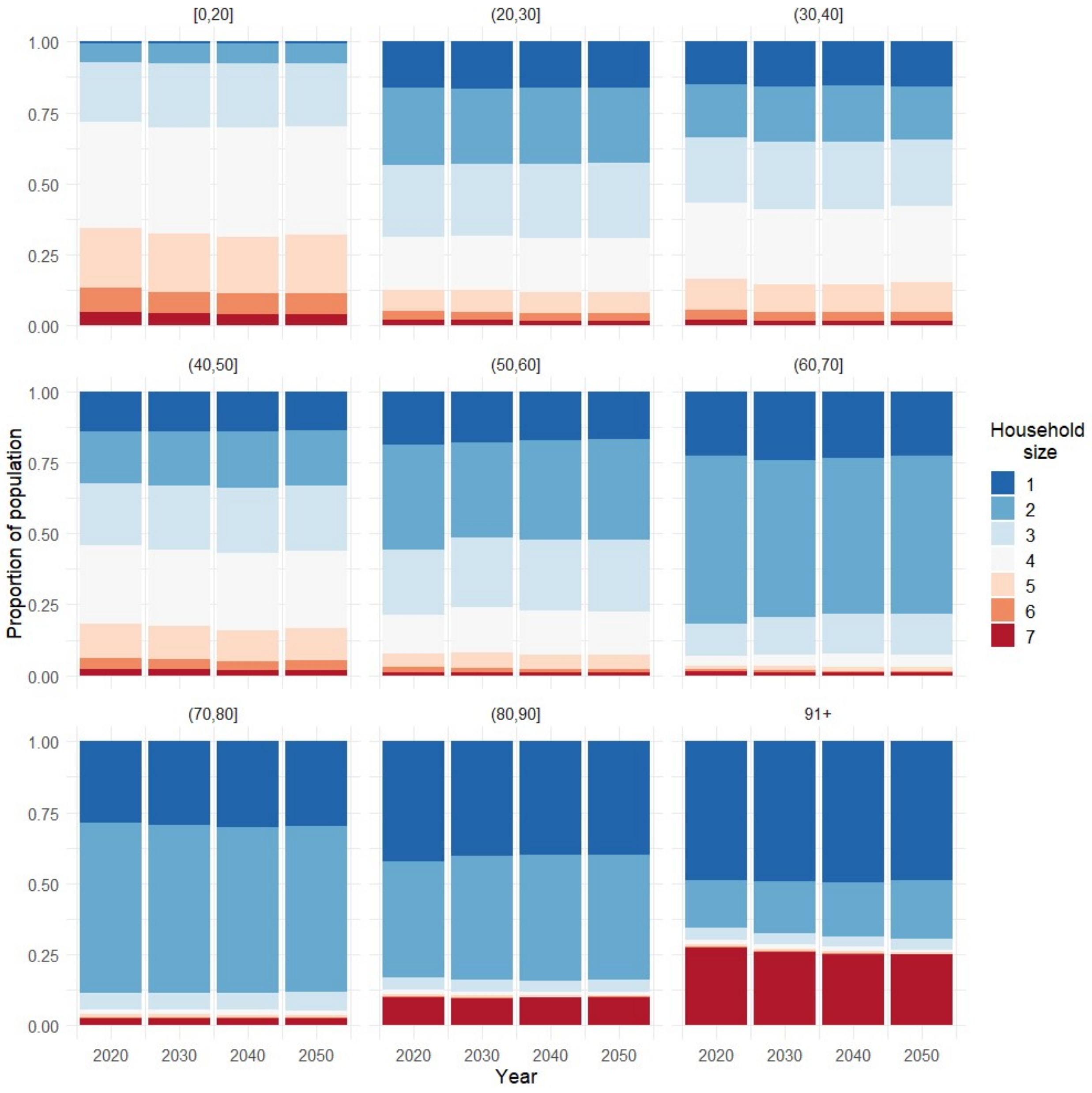

*Figure S3: Household size distribution by age group and year (7=7+).*

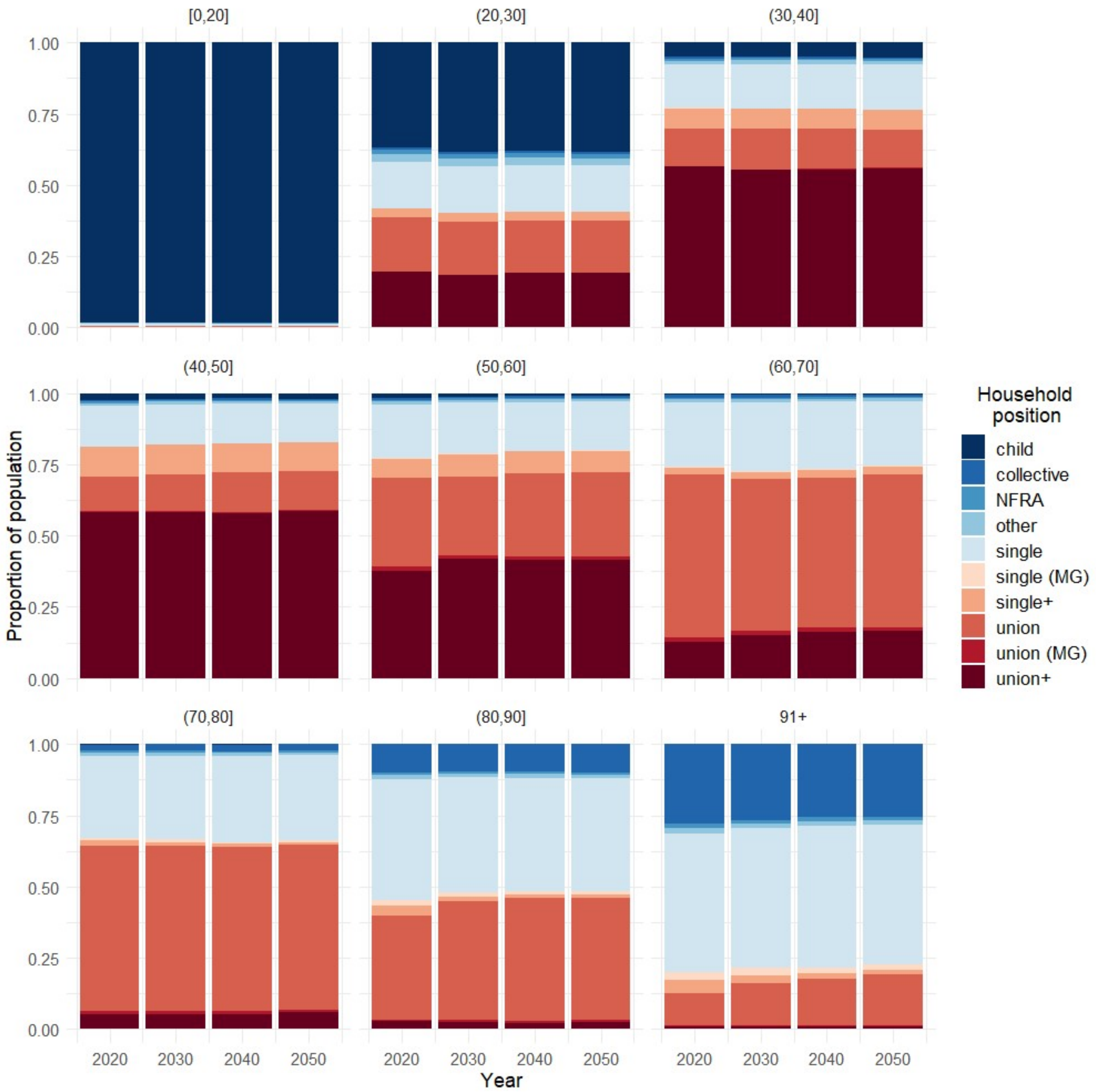

*Figure S4: Household position distribution by age group and year. NFRA: non-family related adult, MG: oldest generation in multi-generational household, single+: single parent, union+: union with child(ren) in household.*

Total fertility rate

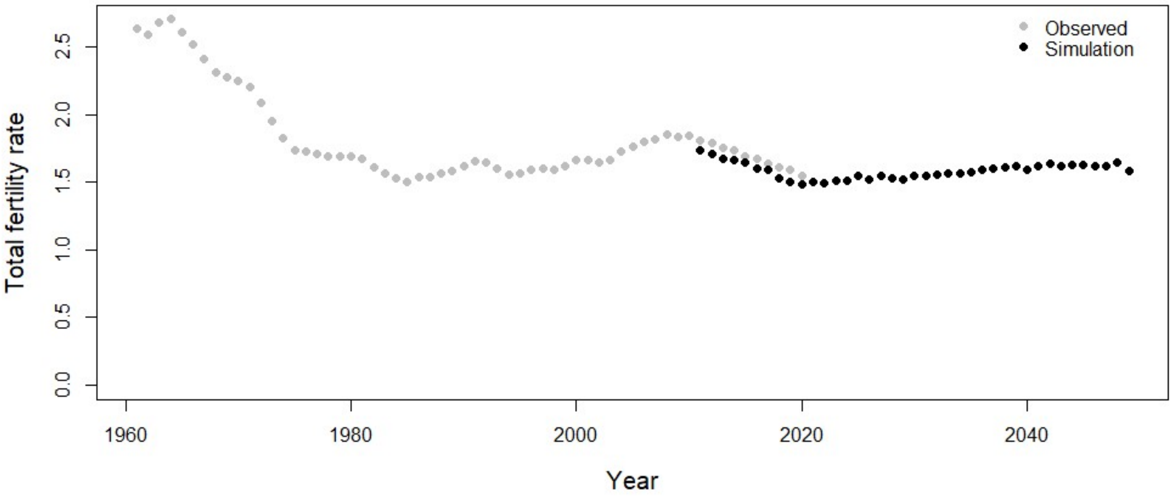

*Figure S5: Total fertility rate. Observed (Statistics Belgium): 1960-2020. Simulation: 2011-2050.*

### Incidence by transmission parameters and scenarios

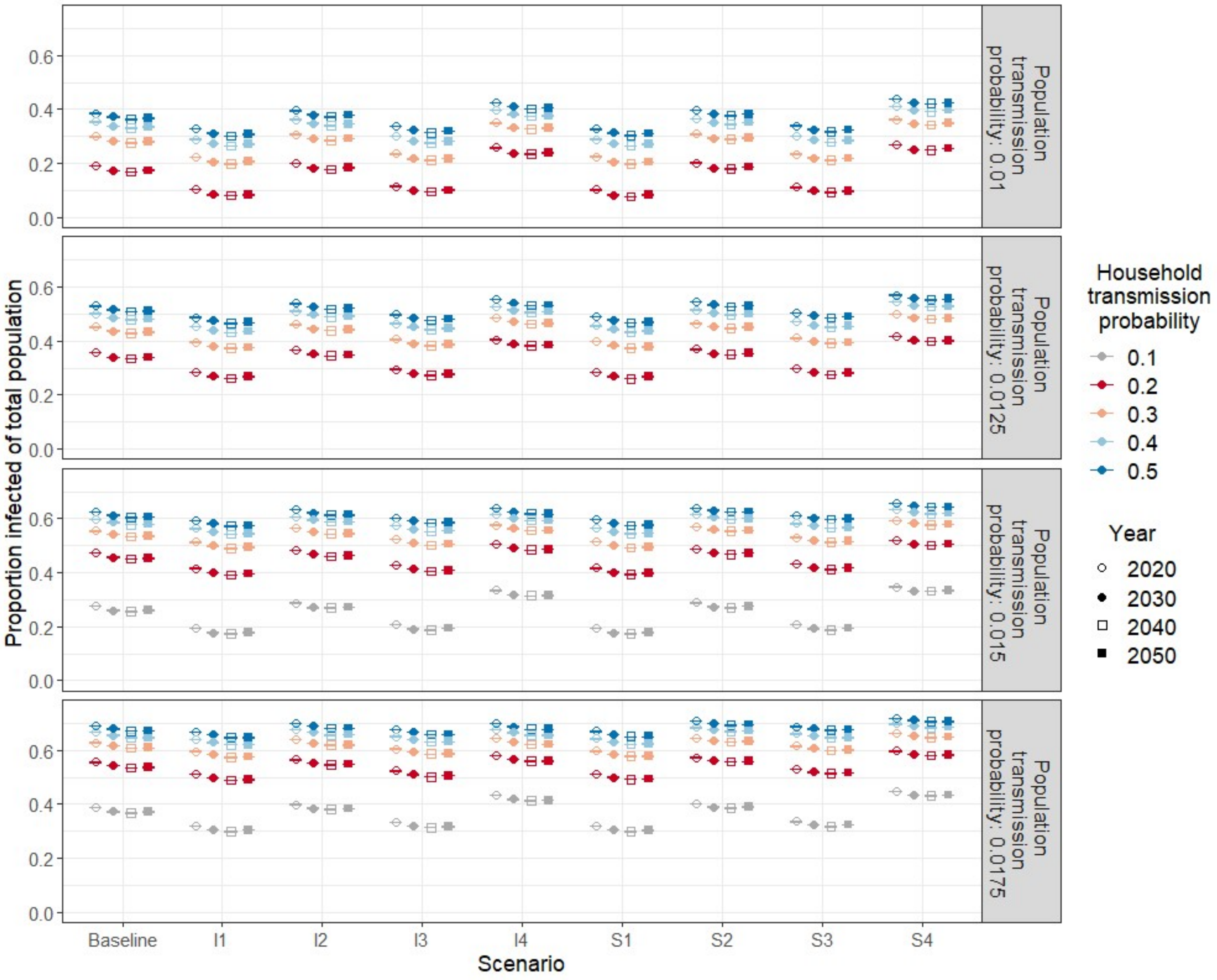

*Figure S6: Mean attack rate in total population with 95% confidence interval for varying transmission parameters ($\beta_h$, $\beta_p$), susceptibility and infectiousness scenarios and simulation year.*

### Threshold parameter $R_*$

We compute the threshold parameter $R_*$ based on Ball et al. (1997). The basic reproduction number $R_0$ is not used because it requires large group sizes, which is not the case for the households in our two-level mixing model. The computation of $R_*$ is based on equation (3.31) in Ball et al. (1997) :

$$R_* = \lambda_G E[T_I] \mu_h^{-1} \sum_{n=1}^{\infty} (1 + \mu_{n-1,1}) n h_n \,, \qquad (1)$$

$$R_* = \mu R_G \quad (2)$$

where $n$ corresponds to household size, $h_n$ is the proportion of households of size $n$ and $\mu_h$ is the mean household size. We compute the average final size in households of size $n$, $(1 + \mu_{n-1,1})$, by starting with one randomly chosen infected individual in each household, which then can pass on the infection to household members, which also can transmit the infection within the household. Meanwhile transmission in the general population is disregarded. Finally, the average final size by household size is calculated and used to compute $\mu = \mu_h^{-1} \sum_{n=1}^{\infty} (1 + \mu_{n-1,1}) n h_n$ (i.e. the average number of household infections). The basic reproduction number in the general population when disregarding household transmission, $R_G = \lambda_G E[T_I]$, is computed by initially infecting one randomly chosen individual in the population. The individual can transmit the infection to others in the population, but the newly infected individuals cannot pass on the infection. The average number of secondary cases is then calculated.

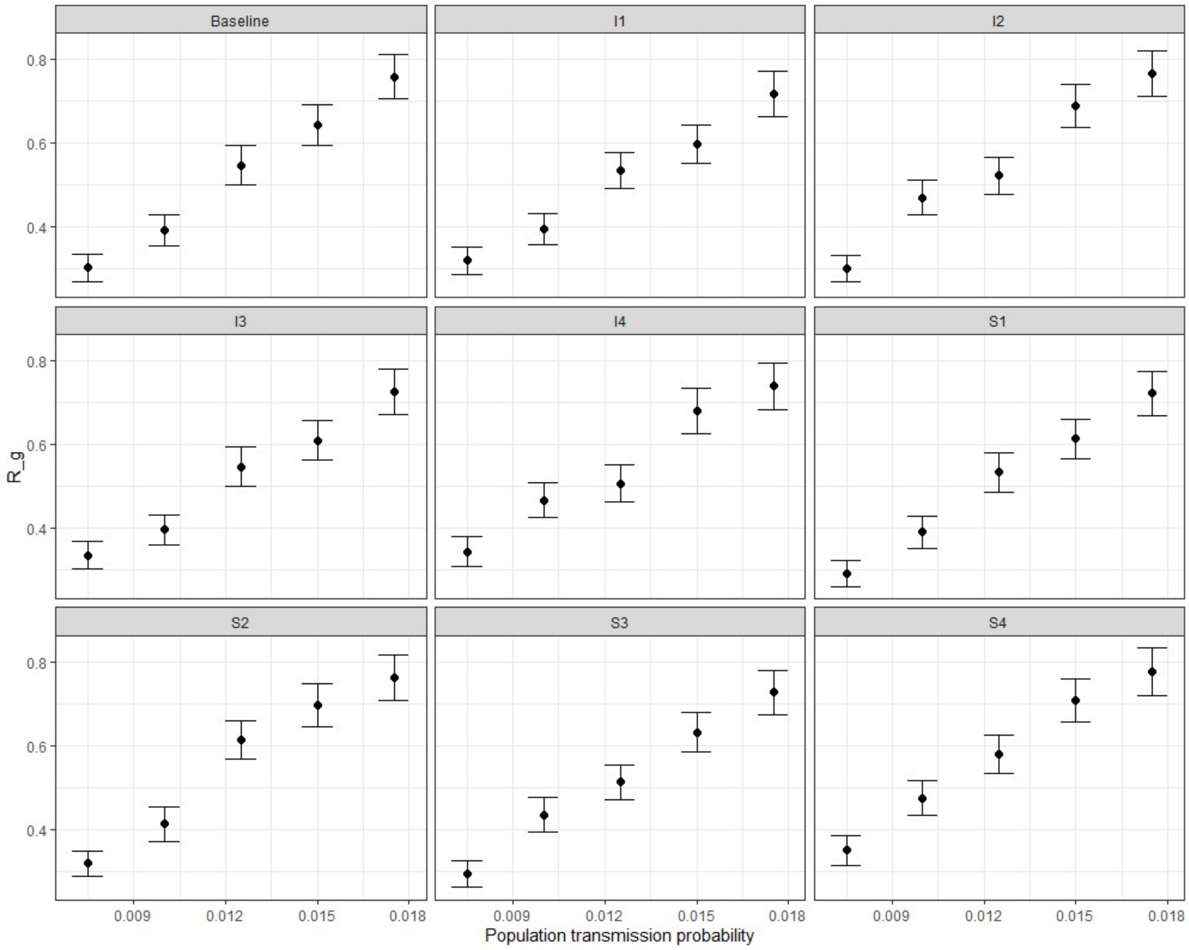

*Figure S7: Estimated $R_G$ in $R_*$ by population transmission probability and scenario (2020 population).*

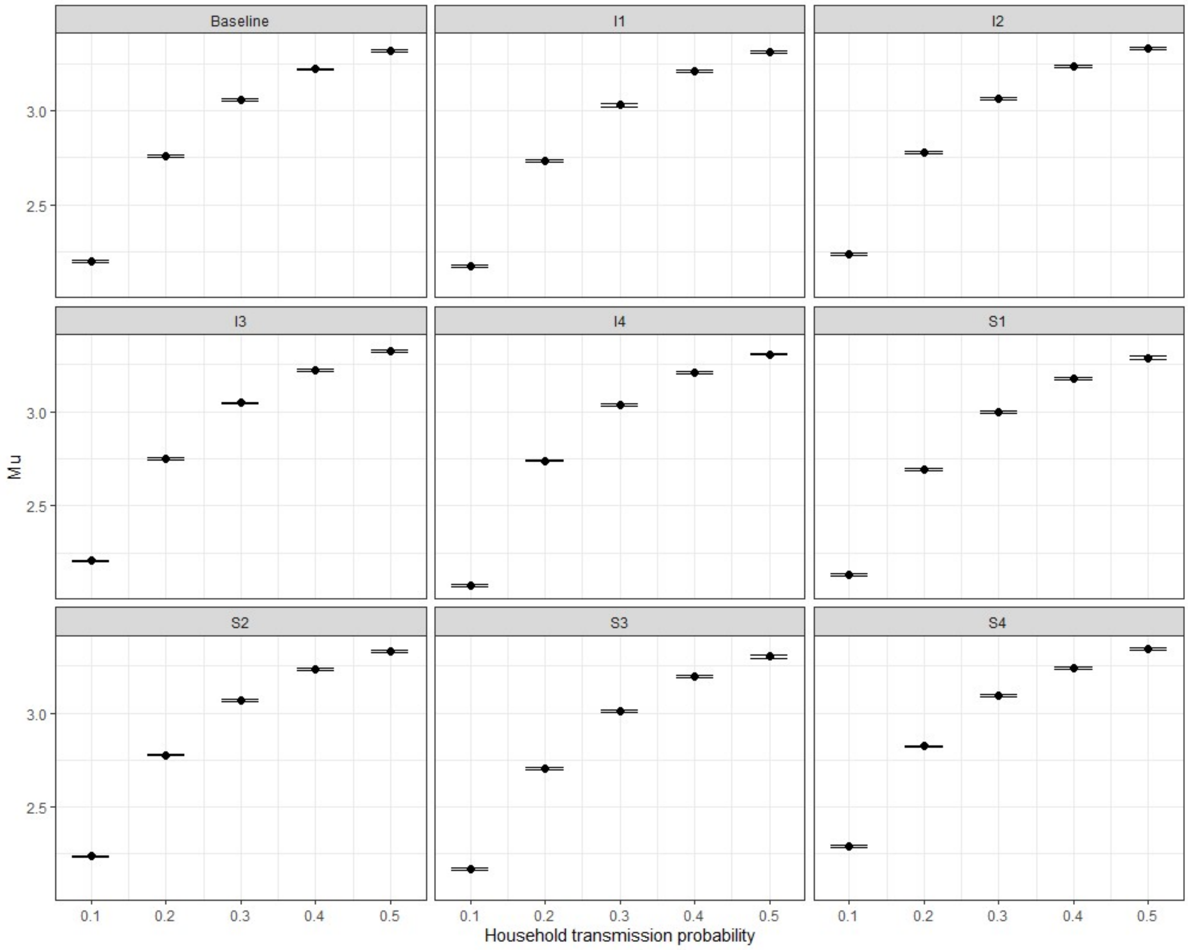

*Figure S8: Estimated μ in $R_*$ by household transmission probability and scenario (2020 population).*

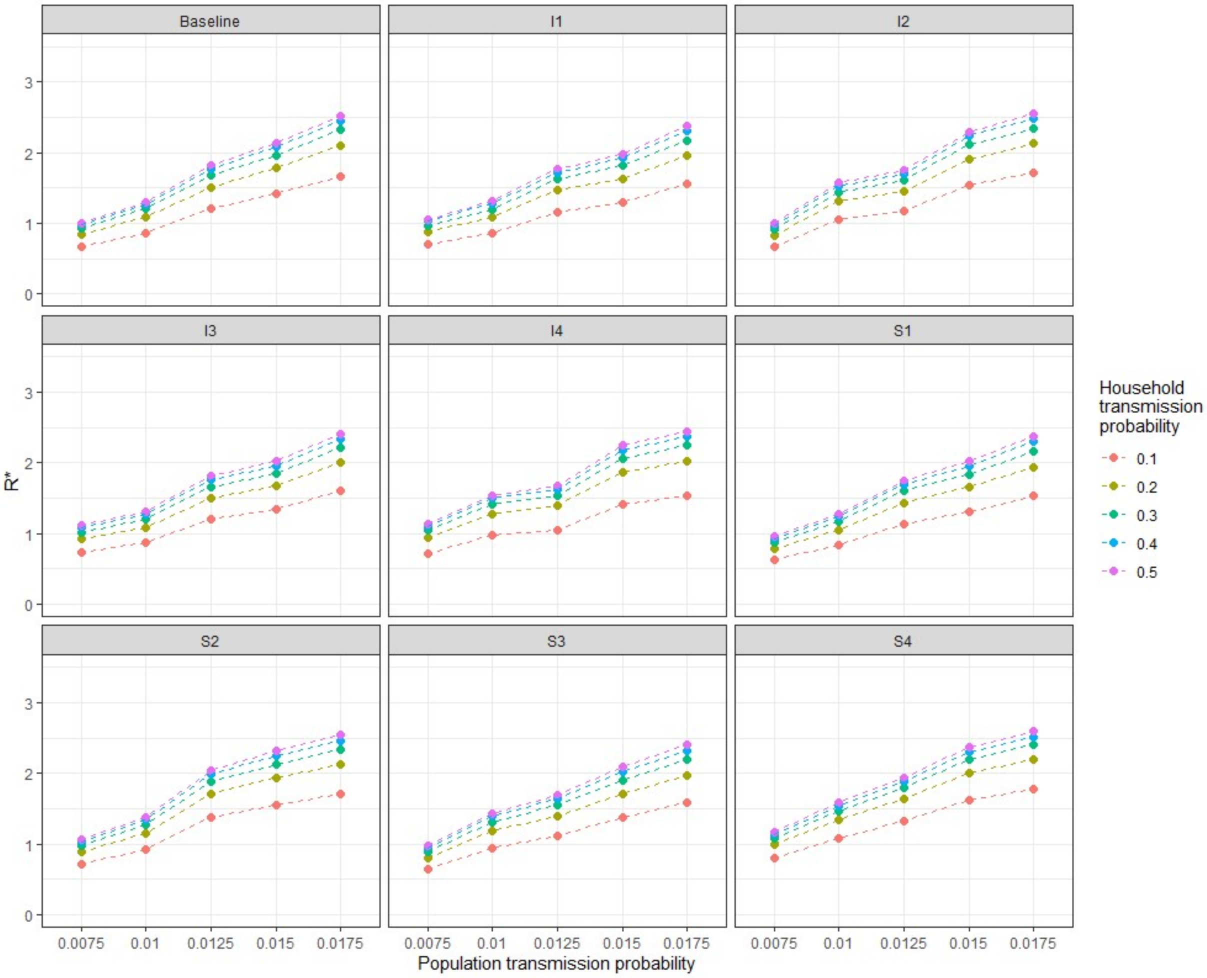

*Figure S9: Estimated $R_*$ by household and population transmission probability and scenario (2020 population).*

### Incidence by household size and type

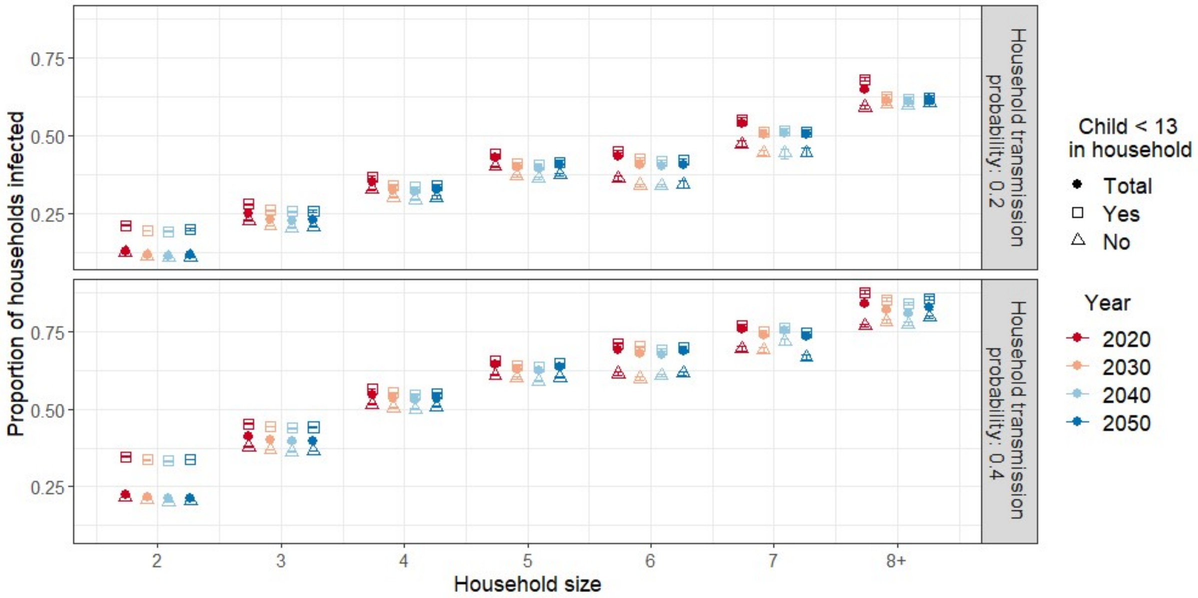

*Figure S10: Mean proportion of households with minimum one infected household member by household size and type in baseline scenario. Population transmission probability: 0.01.*

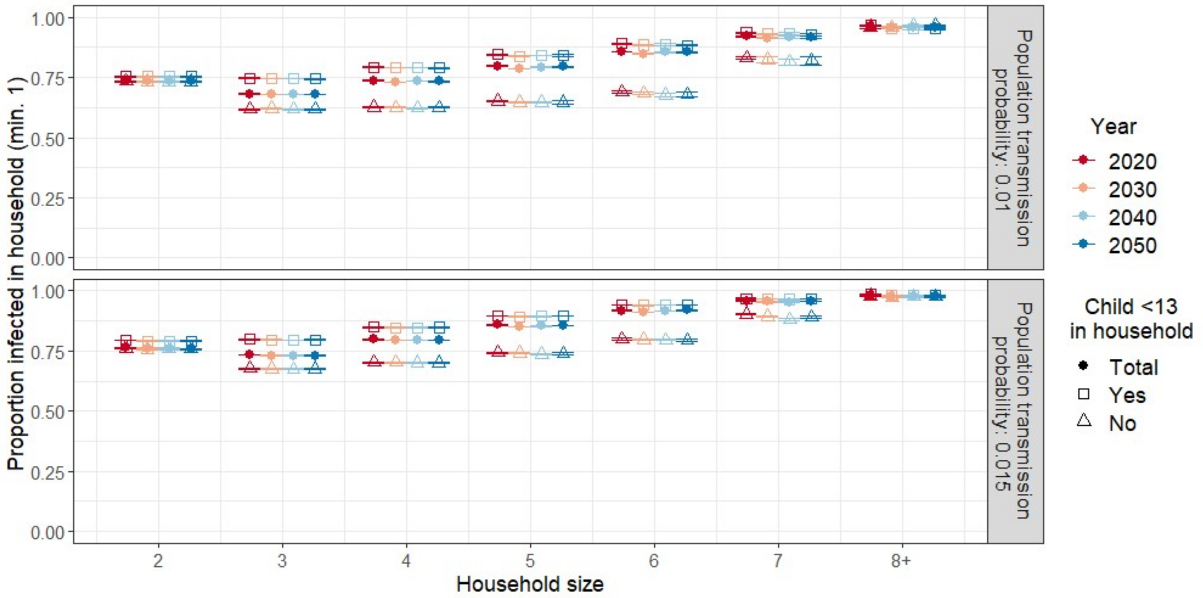

*Figure S11: Mean proportion of household members getting infected by household size and type in baseline scenario. Estimate based on households with minimum one infected individual. Household transmission probability: 0.2.*

### Age-specific incidence relative to baseline and over time

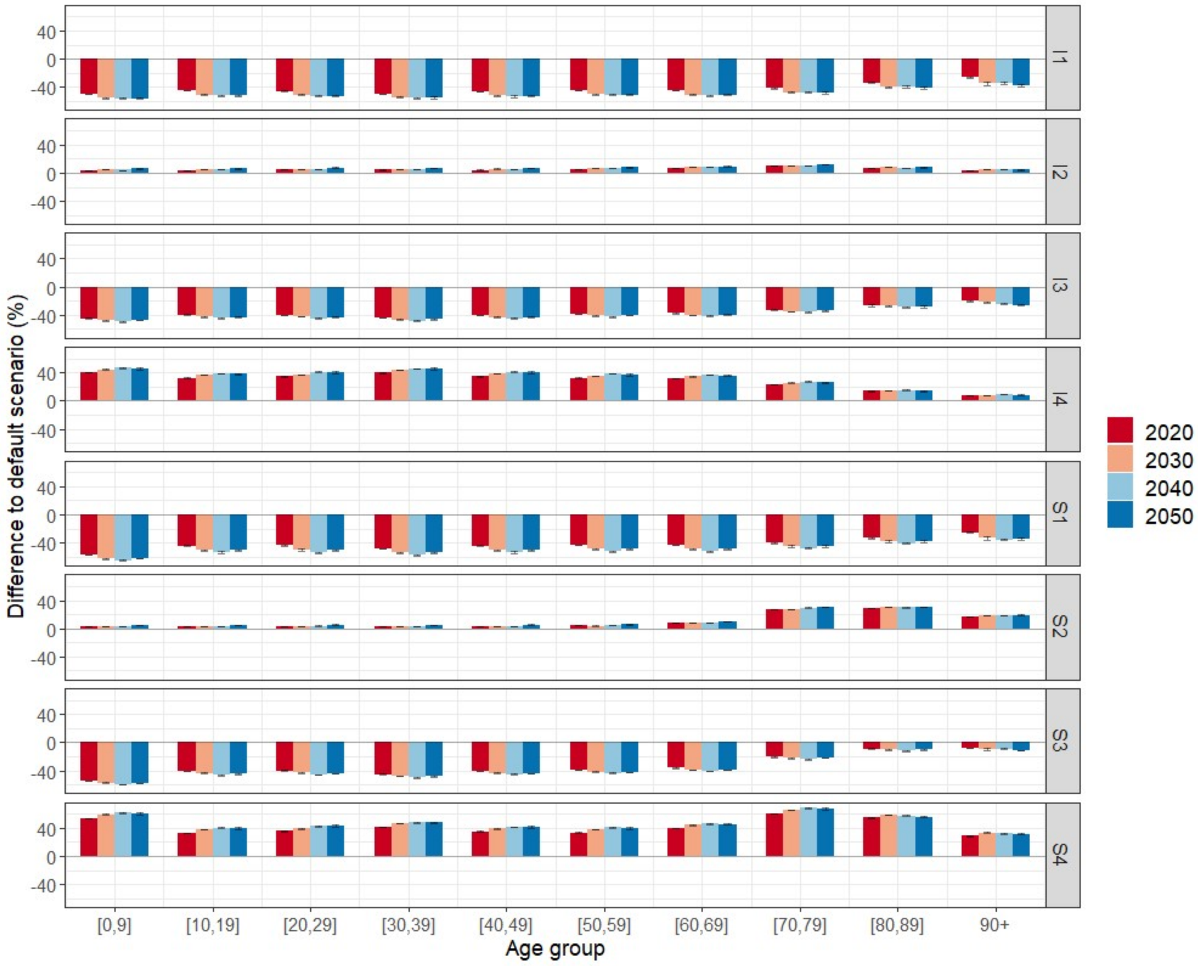

*Figure S12: Percentage difference in age-specific attack rate compared to baseline (equal susceptibility and infectiousness across all ages). Household transmission probability: 0.2, population transmission probability: 0.01.*

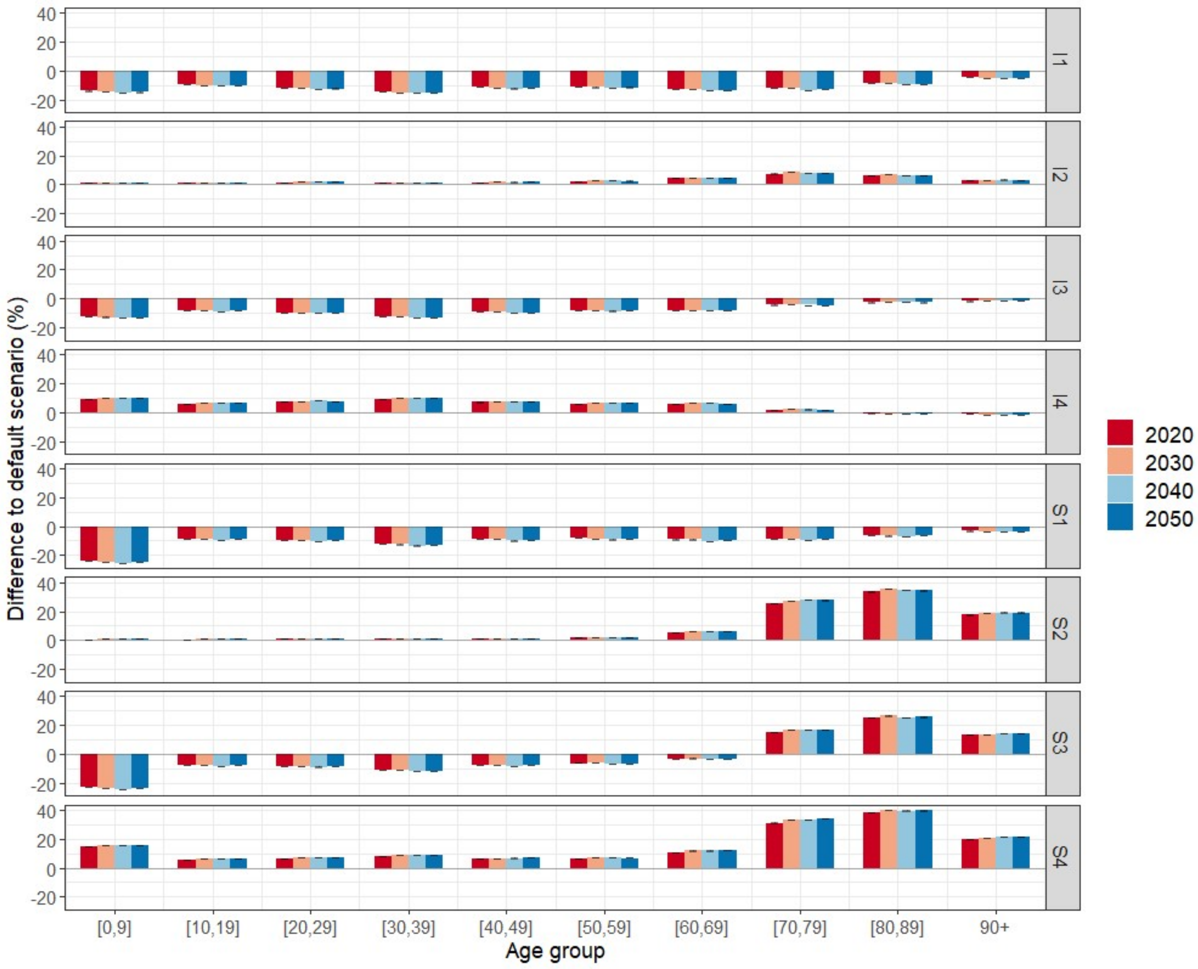

*Figure S13: Percentage difference in age-specific attack rate compared to baseline (equal susceptibility and infectiousness across all ages). Household transmission probability: 0.2, population transmission probability: 0.015.*
